# Personalized Home-Based Cognitive-motor Training using Exergames in Community-Dwelling Older Adults: a Pragmatic Randomized Controlled Trial

**DOI:** 10.1101/2025.05.15.25327487

**Authors:** Julia Seinsche, Eling D de Bruin, Enrico Saibene, Deborah Knechtle, Dino De Bon, Francesco Rizzo, Ilaria Carpinella, Jennifer Rütsche, Maurizio Ferrarin, Ricardo Villa, Sotiria Moza, Eleftheria Giannouli

## Abstract

In an aging population, exergames, a means for simultaneous cognitive-motor training, in a telerehabilitation setting show promise in overcoming treatment accessibility issues. This study aimed to investigate feasibility and effectiveness of a 10-week home-based stepping exergame training program (60 min/week) under remote supervision in older adults as compared to usual care.

An international pragmatic randomized controlled trial (RCT) was conducted in Switzerland, Italy, and Cyprus, including 127 older adults aged ≥60 years, who were randomly allocated to an intervention (n=62) or a control group (n=65).

The adherence rate was 101.4%, the attrition rate 11.8%, and the average exergame enjoyment was rated with 69.1±18.7 out of 100 points. Significant interaction effects were observed in the Go/No-Go test (Q(1, 53.17)=5.44, p=.02) and significant differences between pre- and post-measurements in the intervention group were found when analyzing the 3 study sites separately (ABC (CY, r=.38, p=.05), Go/No-Go (IT, r=3.0, p=.03), Flexi (CH, r=.67, p=.007), TUG (CH, r=.52, p=.04; CY=r.46, p=.02)).

Thus, the step-based exergame intervention appeared to be feasible and safe for older adults, supporting its application in out-patient rehabilitation settings. Additionally, it was effective in improving response inhibition. However, to enhance physical functions, especially in non-acute patients, it may be necessary to increase the training load.

Trial Registration: ClinicalTrials.gov NCT05751551

## Introduction and Objective

In an aging society, the need for rehabilitation increases notably, which in turn poses greater financial and personnel challenges for the healthcare system. Home-based training in form of exergames might be part of the solution to overcome this challenge. Exergames have been defined as video games requiring the player to physically move in order to interactively play those games [1]. Exergames allow the simultaneous training of both, cognitive and physical functions which has shown to be more effective than a consecutive training of those functions [2]–[5]. Previous literature investigating the effects of exergames in community-dwelling older adults has demonstrated improvements in physical [6], [7], cognitive [8]–[10], and psychological functions [11], [12].

Several studies explored the implementation of exergames in a home setting, since many older adults prefer home-based training [13]. Key advantages of home-based training include a higher accessibility compared to traditional supervised exercising coupled with an increased enjoyment and motivation/engagement through gamification [14]–[16]. Both, in turn, seem to contribute to an improved adherence to exercise programs in older adults [16] as well as a higher cost-effectiveness [17], [18].

However, findings regarding the effects of home-based exergame training on physical and cognitive functions in older adults are inconsistent. Many previous studies have utilized commercial exergame systems, which were not developed specifically for older adults, or did not consider taking user opinions into account when designing the exergames’ software and hardware. Consequently, many existing exergame systems have failed to meet older adults’ requirements such as simplified user interface, personalized training, and detailed instructions [16], [19].

This study aimed to test the feasibility and effectiveness of a home-based exergame training program which was specifically designed for rehabilitation purposes using a user-centered approach in older adults [19], [20].

## Methods

### Trial design

This trial is an international, pragmatic, two-arm pilot randomized controlled trial (RCT). It was conducted in three countries: (1) Switzerland (VAMED Rehabilitation Center Zurich Seefeld, Zurich), (2) Italy (Rehabilitation units of IRCCS Fondazione Don Carlo Gnocchi, Milan), and (3) Cyprus (Materia Unit, Materia Group, Nicosia). The study protocol was approved by all local ethical committees, including the Cantonal ethics committee in Zurich, Switzerland (2022-01746); the Don Carlo Gnocchi Foundation ethics committee in Italy (06_16/12/2022); and the Cyprus National Bioethics Committee (EEBK EIT 2021 51). It has been registered at ClinicalTrials.gov (NCT05751551) and the study protocol is published [21]. The reporting of this trial is based on the CONSORT checklist [22] and its extensions for pilot and feasibility trials [22] and for pragmatic trials [23].

### Participants

Between January and September 2023, community-dwelling older adults were recruited via convenience sampling, advertisements in newspapers and -letters, flyers, announcements, and presentations in various senior organizations, such as the University of the Third Age of the University of Zurich, and via a home care provider (Spitex Zurich). Eligibility was tested during an initial screening phone call and a subsequent personal meeting. Major inclusion criteria were (1) age ≥60 years (2) prescription for rehabilitation (as inpatient or outpatient) within the past 6 months, (3) Mini-Mental State Examination (MMSE) score ≥24, and (4) ability to stand independently for at least 2 minutes. Major exclusion criteria were (1) permanent residence in a nursing home, (2) mobility limitations, cognitive limitations, or comorbidities impairing the ability to perform step-based, cognitive-motor training (3) severe sensory impairments, (4) previous or acute major psychiatric illness (5) participation in another clinical trial, and (6) absence from home of >2 weeks during the trial period. Before testing for eligibility and before collection of baseline data, informed consent had to be obtained.

### Randomization and blinding

Following the completion of all baseline assessments, participants were randomly allocated to one of 2 groups (an intervention group (IG) or a passive control group (CG)) with a 1:1 allocation ratio using permuted block randomization. The allocation list, based on the random generation of blocks of 4, 6, and 8, was generated by the web-based randomization tool Sealed Envelope [24] and uploaded on REDCap (an electronic data capture tool [25], [26]) by a study investigator not involved in the enrollment, baseline assessment, and randomization of participants. No other investigator had access to the randomization list, ensuring allocation concealment. The randomization was then executed in REDCap subsequent to the baseline assessments.

Given the nature of the intervention and the passive control group, neither participants nor the investigators could be blinded. Additionally, due to limited personnel resources, the investigators responsible for supervising the interventions were also conducting the measurements, which is why outcome assessors were not blinded either.

### Interventions

The detailed intervention description is available in the study protocol [21]. Participants in the IG initially engaged in a 2-week supervised exergame training to get familiarized with the exergame device system – the Dividat Senso (Dividat AG, 8834 Schindellegi Feusisberg, Switzerland). The Dividat Senso is a stationary stepping platform consisting of 5 pressure sensitive plates connected to a screen displaying exergame stimuli and part of the COCARE system. Subsequently, participants proceeded to a 10-week unsupervised exergame training at home using the Dividat Senso Flex - a portable pressure-sensitive mat with the same functionalities as the Dividat Senso. Participants were instructed to train for a total of 60 minutes per week, preferably in 3 training sessions. In both intervention periods, supervision was provided by trained health science master students and physiotherapists.

A total of 11 step-based exergames were implemented, designed to target dynamic balance and cognitive functions including attention (selective and divided), action planning, cognitive flexibility, inhibition, visuo-spatial orientation, and working memory. Additionally, 2 exergames were specifically implemented to train endurance through walking-on-spot, and 4 weight shifting games targeting static balance were incorporated. More detailed game descriptions can be found in Supplementary Table 4.

The training plan encompassed 15 levels for each trained function (cognition, balance, and endurance), defined by game selection and duration. Initial levels were determined based on several motor and cognitive-motor assessments provided by the Dividat Senso.

The home-based intervention started with an initial home-visit to install the Senso Flex and further on, biweekly phone calls were conducted. Unlike announced in the protocol, additional home visits were only arranged in case of technical issues or other difficulties, as the regular phone calls were sufficient for addressing other concerns. Both groups continued receiving their usual care such as physiotherapy or ergotherapy throughout the study participation period.

### Monitoring

Participants’ training durations and performance were recorded using the system’s online proprietary software (Dividat Manager). Participants who did not train at all for an entire week were contacted via telephone to identify potential issues or barriers and, if necessary, to adapt the training plan.

### Outcomes and outcome measures

Prior to the study start, detailed standard operating procedures (SOPs) were developed including detailed information about the order of assessments, the precise instructions which were to be given as well as the exact execution of each assessment. To ensure full understanding, these SOPs were provided in written form and presented and discussed comprehensively in a personal meeting.

#### Primary outcomes

The primary outcome of this study was feasibility which was determined by the following measures: (1) adherence to the training program, defined as the percentage of actual training duration and number of trainings per week in relation to the recommendations; (2) attrition rate, defined as the percentage of dropouts; and (3) exergame enjoyment measured with the Exergame Enjoyment Questionnaire (EEQ) [27]. The following criteria should have been met for the intervention to be deemed feasible: ≥70% adherence rate and attrition rate of ≤20%.

#### Secondary outcomes

Secondary feasibility outcomes encompassed the National Aeronautics and Space Administration Task Load Index (NASA-TLX) [28] to assess the subjective workload during training, and participants’ willingness to continue a similar home-based exergame training program after study completion (measured on a 5-point Likert scale). Additionally, number and types of additional instructions and help requests were noted. These secondary feasibility outcomes were assessed at the end of the intervention period.

Cognitive-motor functions were evaluated through 3 step-based cognitive-motor assessments using the Dividat Senso: (1) the Reaction Time Test (RTT) measuring psychomotor speed, (2) the Go/No-Go Test to assess selective attention and inhibition, and (3) the Flexibility Test as a measure of mental flexibility. Furthermore, two balance tests were conducted on the Dividat Senso: (1) the Sway Test measuring postural sway while standing still for 30 seconds, and (2) the Coordinated Stability Test evaluating functional stability by directing the participants to follow a given figure on the screen by shifting their center of pressure (COP). To investigate mobility and functional balance, and dual-task ability, the Timed up-and-go (TUG) [29] and timed-up-and-go dual task (TUG-DT) were used. For the latter, a secondary cognitive task was added, which in this study meant counting backward from 90, subtracting 7 successively. Additionally, leg power was assessed using the 30-second Sit-to-Stand Test [30]. Psychological outcomes comprised two questions about the perceived quality of life and health satisfaction, as well as balance confidence measured with the Activities-specific Balance Confidence (ABC) Scale [31]. All secondary cognitive, physical, and psychological outcomes were assessed before and after the home-based intervention period.

#### Other outcomes

Data concerning participants’ usual care as well as their daily cognitive and physical activities were collected through biweekly phone calls. Furthermore, the following demographic information was recorded: age, sex, years of education, Mini Mental State Examination (MMSE) total score, living situation, Body Mass Index (BMI), reason for therapy prescription, number of falls, and comorbidities.

### Sample size

Sample size considerations were based on recommendations provided by Whitehead et al. [32]according to which, in pilot clinical trials, targeting an extra small standardized effect size (≤0.1) with 90% power, a minimum of 75 participants per group is necessary. Anticipating a dropout rate of 20%, we intended to enrol 180 participants (n=90 per group; n=60 per trial site).

### Statistical methods

Descriptive statistics as well as the analysis of feasibility measures were conducted with SPSS (Version 28), whereas all analyses of effectiveness were performed with R Studio (Version 2021.09.0).

For comparing descriptive statistics between two groups, independent, 2-tailed t-tests (interval data), Chi-Square tests (binominal variables), and Mann-Whitney-U tests (ordinal data) were conducted. When comparing descriptive statistics of the three study sites, one-way ANOVAs with Tukey’s post hoc tests were conducted for interval data. As a robust alternative, in case of heterogeneity of variances, Welch test and Games-Howell as post hoc test were interpreted. Chi-Square tests, and Kruskal Wallis tests were used for comparing binominal, respectively ordinal data.

Concerning the measures of effectiveness, robust mixed ANOVAs (R-function bwtrim() [33]) were applied to investigate within and between group effects as well as interaction effects. This deviates from the study protocol according to which a linear mixed model was planned to create and analyze. However, normal distribution and homoscedasticity of residuals were not given in most variables which is why a robust analysis was required. Ordinal data was analyzed with a non-parametric ANOVA using the package “nparLD” [34]. To further explore the results of the robust mixed ANOVAs, Wilcoxon signed rank tests were used to analyze within-group differences and to achieve effect sizes.

Per-protocol analyses were conducted when investigating the effectiveness of the training program including data of only those participants who underwent the whole study procedure, with a mean training adherence of at least 70% during the home-based training. The results were compared with an analysis of all participants participating in the follow-up assessments. All statistical tests were two-sided, with statistical significance defined as p < 0.05.

## Results

In total, 145 people were screened for eligibility between October 2022 and November 2023. Only one person was found ineligible, resulting in the inclusion of 144 participants in the study who were randomized to either the intervention group (n=73) or the control group (n=71). Ultimately, 17 participants withdrew from the study. In the intervention group, 3 participants withdrew immediately after T1 without starting any training, 6 during the supervised training period or shortly after T2, and 2 during the home-based training period. Thus, 127 participants were included in the analyses (Switzerland n=21, Italy n=55, Cyprus n=51) (Figure 1). Participants who withdrew after being enrolled in the study did not differ significantly from those who completed the study in terms of age (p=.72), sex distribution (p=.50), years of education (p=.27), MMSE score (p=.24), or Comorbidity index (p=.87).

**FIGURE 1.**
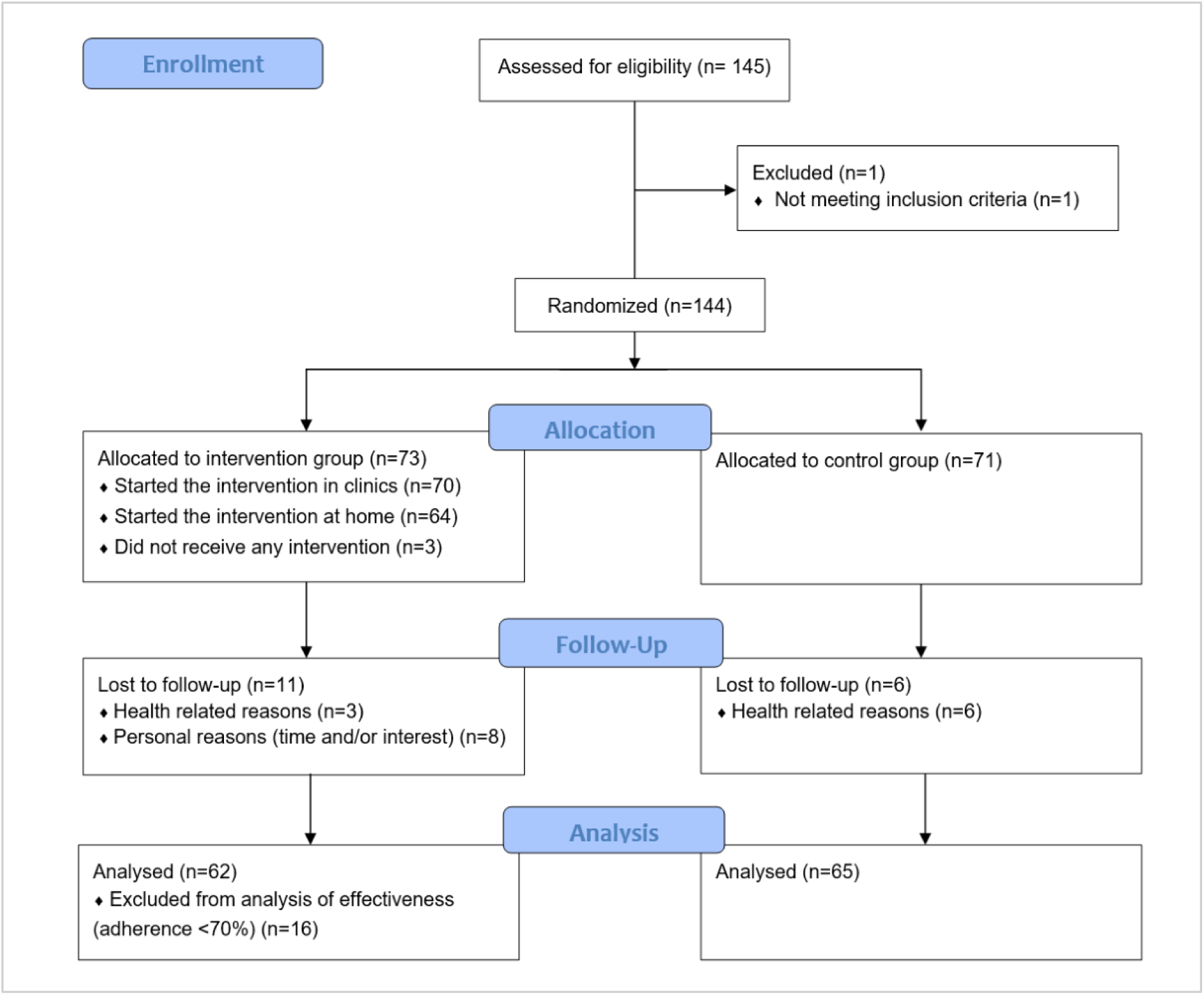
CONSORT Flow diagram. Number of participants assessed for eligibility, randomized, and included in the analysis

### Demographics

Participants who were included in the final analysis had a mean age of 73.0±7.3 years and 78% were female. Reasons for prescription of rehabilitation were primarily orthopedic (n=57), followed by general preventive treatments or maintenance of motor and/or cognitive functions, respectively (stimulation therapy) (n=29), and neurological conditions (n=32) of which 5 had Mild Cognitive Impairment (MCI) and/or memory problems, 9 a stroke, 11 Parkinson’s disease, and 2 Multiple Sclerosis. The remaining participants were diagnosed with internal diseases (n=4). Table 1 displays further demographic characteristics of the intervention group (IG) and the control group (CG) and Supplementary Table 1 provides a comparison of demographic characteristics of the different trial sites. IG and CG did not differ in terms of age (p=.49), sex distribution (p=.12), MMSE score (p=.57), BMI (p=.51), years of education (p=.14), or the Charlson Comorbidity Index (p=.22), which is why no adjustments were made for the analyses.

**TABLE 1.**
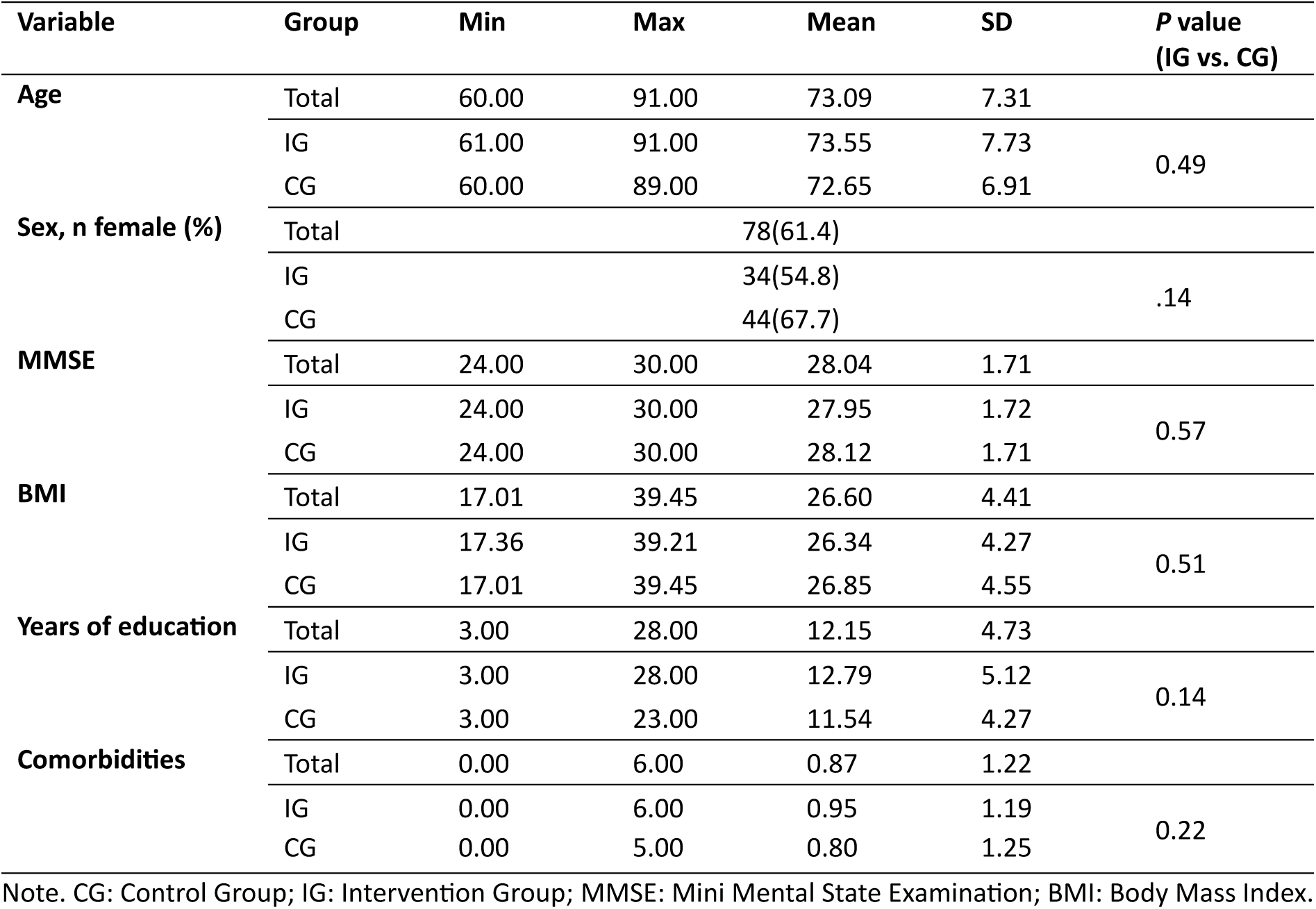
Demographics (total sample n=127, IG: 62, CG: 65))

However, comparing study sites, statistically significant differences were found for age (F(2,124)=3.37, p=.04, η²=.052), years of education (F(2,124)=7.51, p<.001, η²=.108), MMSE (Welch’s F(2,65.44)=12.28, p<.001, η²=.129), and the Charlson Comorbidity Index (H(2)=39.704, p<.001, r=-.11). Post hoc tests and pairwise comparisons, respectively, did not identify significant differences in terms of age, but showed that participants from Switzerland and Italy had a higher number of years of education than Cyprus (4.32, 95% CI[1.55-7.09], p=.001, and 2.18, 95% CI[0.12-4.26], p=.04), a slightly but significantly higher MMSE score (1.65, 95% CI [0.84-2.45], p<.001, and 0.9, 95% CI [0.18-1.73], p=.01), and a higher Comorbidity index (p<.001, z=5.81, r=z/sqrt(72)=.68; p<.001, z=4.51, r=z/sqrt(106)=.44). Sex distribution (p=.20) and BMI (p=.10) did not differ between study sites.

Concerning the other demographic factors, 87 (68.5%) participants reported living with another person and 40 (31.5%) were living alone. The living situation differed significantly between groups (IG: 21% living alone, CG: 41.5%, p=.013) and between study sites (p=.02) with the highest percentage of people living alone in Switzerland (52.4%), followed by Cyprus (35.3%), and Italy (20%). Furthermore, 95 participants (74.8%) were classified as non-fallers, whereas 32 (25.2%) experienced at least one fall within the past 12 months (IG: 30.6% faller, CG: 20%, no significant group difference: p=.167). Significant site differences in fall status were found (p<.001) with highest percentage of fallers in Italy (36.4%), followed by Switzerland (33.3%) and Cyprus (9.8%).

### Feasibility

#### Attrition and adherence

17 participants withdrew from the study, which equals an attrition rate of 11.81%. Concerning adherence, over the course of the whole intervention period, the participants trained for an average of 60.8 minutes per week, which equals an adherence rate of 101.43%. Thereby, participants from Switzerland showed the highest adherence to the training program, followed by Italy and Cyprus (Table 2). However, these differences were not statistically significant (p=.131).

**TABLE 2.** Adherence [%] (training time as percentage of recommended training time)

| | Min | Max | Mean (SD) | Median | P value, $\eta^2$ |
| --- | --- | --- | --- | --- | --- |
| Total (n=62) | 11.25 | 337.67 | 101.43 (57.8) | 87.42 |  |
| Switzerland (n=9) | 52.17 | 269.17 | 117.82(62.69) | 106.75 |  |
| Italy (n=30) | 11.25 | 227.05 | 111.12(54.18) | 98.92 | $p=.131$, $\eta^2=.07$ |
| Cyprus (n=23) | 26.67 | 337.67 | 82.36(57.94) | 73.33 |  |

**TABLE 3.** Exergame Enjoyment.

|  | Min | Max | Mean (SD) | Median | P value, effect size |
| --- | --- | --- | --- | --- | --- |
| Total (n=62) | 25 | 93 | 69.13(18.72) | 73.0 |  |
| Switzerland (n=9) | 70 | 85 | 78.11(6.70) | 80.0 | $P<.001$, $\eta^2=.381$ |
| Italy (n=30) | 25 | 79 | 57.33(20.10) | 68.0 |  |
| Cyprus (n=23) | 66 | 93 | 81.0(7.26) | 82.0 |  |
| High adherer (n=46) | 25 | 93 | 66.37(20.06) | 72.0 | $P=.012$, $d = 1.74$. |
| Low adherer (n=16) | 46 | 93 | 77.06(11.23) | 78.0 |  |

Figure 2 illustrates the temporal change of adherence rates. It is apparent that across all trial sites, participants’ adherence decreased slightly from Week 1 to Week 10, albeit to differing degrees. The average adherence rate, however, maintained relatively stable until Week 8 before declining as well. Nonetheless, throughout all time points and across all trial sites, the mean adherence rate consistently exceeded the threshold of 70% which has been defined as “being adherent to the training” in our study protocol [21] similar as in other previous research [35], which indicates a good feasibility of the training program.

**FIGURE 2.**
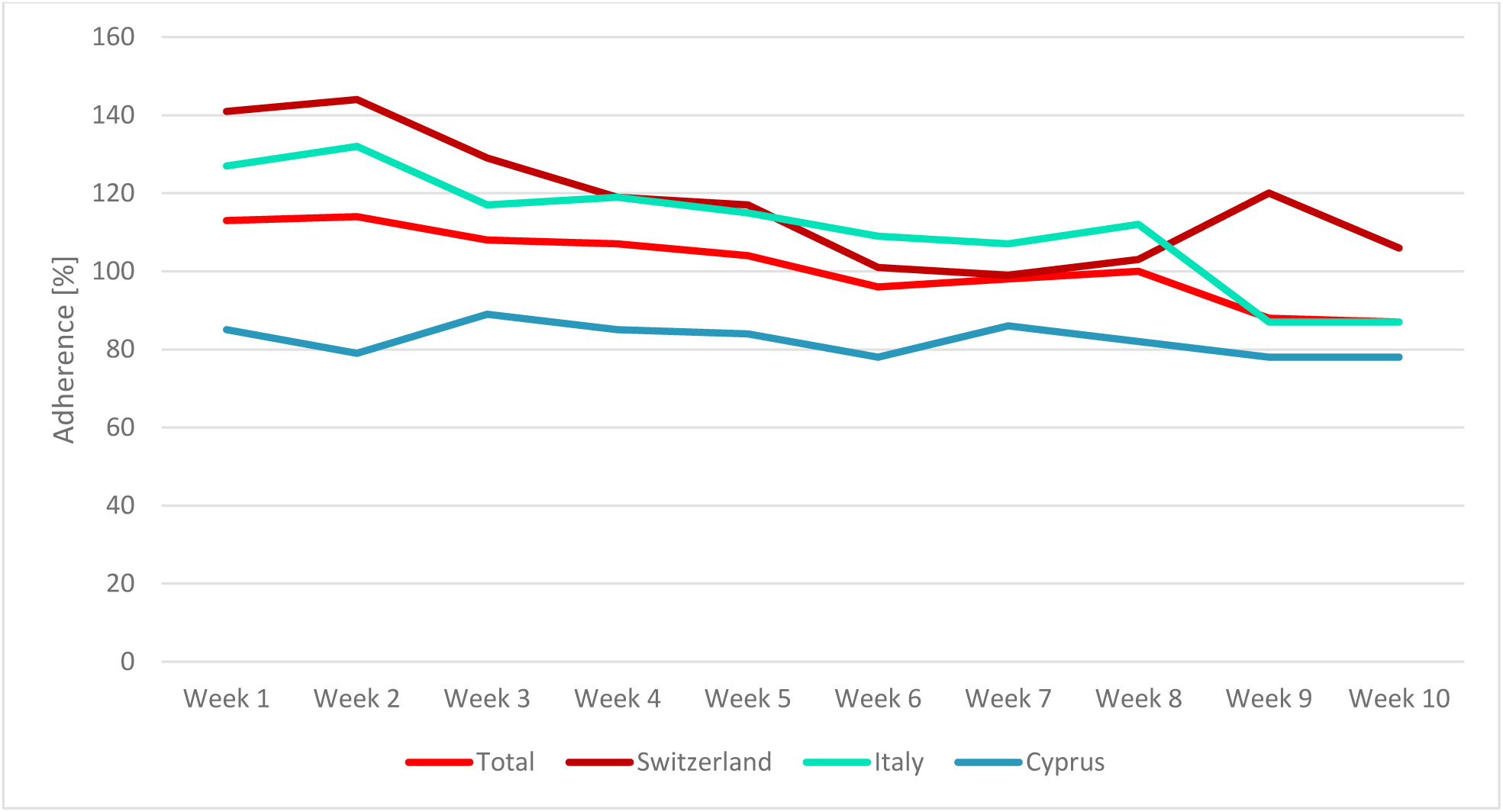
Adherence rates over time (overall and divided by study site)

#### High and Low adherers

As announced in the study protocol, participants training for less than 70% of the recommended training time, were excluded from the analyses of effectiveness. This was the case in 16 participants (Switzerland: n=1, Italy: n=5, Cyprus: n=10). Those participants had an average adherence rate of 49.71 ± 19.0%, whereas in high adherers with an average adherence rate of ≥70%, the rate was 119.41±56.0% (Figure 3).

**FIGURE 3.**
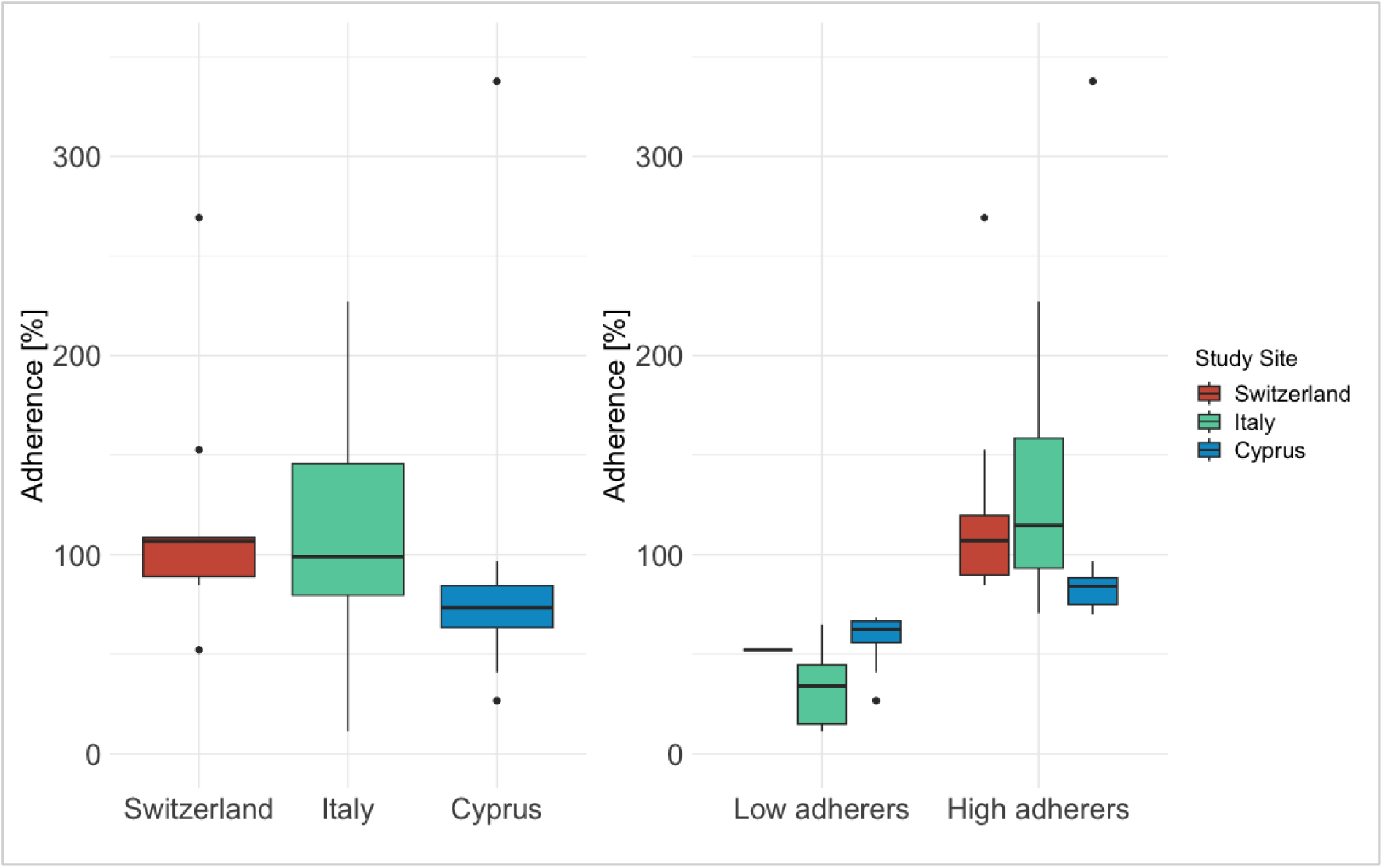
Boxplots displaying the distribution of adherence rates divided by study site and adherence group (low adherers: adherence rate < 70%). All boxplots show the median line, the 3rd and 5th quartile, and the interquartile range (IQR), and the whiskers indicate the range of all values within 1.5 times of the IQR. Values outside this range are considered outliers and plotted as dots.

High and low adherers did not differ in terms of any demographic factor (age (*t*(60) = 1.253, Cohen’s *d=.*364 [95% CI: -.210-.935], p =.215), MMSE score ((*t*(60) = -.879, Cohen’s *d*=-.255 [95% CI: -.825-.317], *p* =.383), years of education (*t*(60) = .358, Cohen’s d=.104 [95% CI: -.466-.673], p =.722), gender (χ²(1) = 1.685, *p* = .194, φ = .165), living situation (χ²(1) = .212, *p* = .646 φ = -.058), fall status (χ²(1) = .323, *p* = .570, φ = .072), or comorbidities (*U* = 368.00, *p* >.90)).

Finally, as in the whole sample, adherence rates did not significantly differ between study sites neither in high (F(2,43)=.913 p=.41, η²=.04), nor in low adherers (F(2,13)=3.303, p=.069 η²=.34).

#### Exergame Enjoyment

Participants rated their enjoyment with an average EEQ score of 69.13±18.72, whereby statistically significant site differences were detected (Welch’s F(2,26.97)=17.376, p<.001, η²=.381), with participants from Cyprus enjoying the training the most, followed by those from Switzerland. Comparing high and low adherers, a significant difference was found as well, however, contrary to expectations, low adherers indicated higher enjoyment levels compared with high adherers (Table3, Supplementary Figure 1).

#### NASA-TLX

On average, participants perceived the training as low to moderately demanding. The mean total raw NASA-TLX score was 40.13±15.84 on a scale of 0–100, whereby the highest subjective workload was reported for the items “effort” (47.74±23.18) and “mental demand” (47.26±28.42), followed closely by “physical demand” (41.69±27.58), and “temporal demand” (40.97±22.94). Furthermore, on average, the participants’ frustration was low (23.31±21.13), and participants rated their performance with a mean score of 39.84±5.25.07. Supplementary Figure 2 illustrates the NASA TLX scores divided by study sites.

Significant site differences were found in the items “mental Demand” (Italy vs. Cyprus: r=.42, p=.006), “physical Demand” (Italy vs. Cyprus: r=.55, p<.001), and “frustration” (Switzerland vs. Cyprus: r=.63, p=.001, Italy-Cyprus: r=.40, p=.012). High and low adherers did not significantly differ in any item (p=.119-.556).

#### Help requests (number and kind)

Over the whole home-based training period, participants directly sought assistance 15 times. Among those help requests, 8 were based on technical problems with the system (computer or mat), 6 participants were logged out (one participant twice) and needed help to re-access the training environment, and one participant asked for additional game instructions. In some participants, the latter occurred more frequently. However, these participants had acquired the ability to independently manage it themselves by that point.

In addition to those direct help requests, some participants expressed some criticism without a clear help request. Common criticisms included dissatisfaction with the low step detection sensitivity of the mat - in particular during balance games (6 participants, 10 complaints), the training plan, especially in terms of the amount or duration of games and the slow progression speed (n=3), and some games (n=22). Regarding the games, most criticism was directed towards the game “Ski” for its high speed and the high number of obstacles (n=9) and towards “Rockets” for not being sufficiently entertaining (n=6).

#### Willingness to continue a similar home-based exergame training program

Overall, most participants (n=43, 69.3%) indicated a willingness to continue the training program (hypothetical question), while some remained neutral (n=12, 19.4%), and only a few declined (n=7, 11.29%). Consequently, the median agreement among all participants was 4 points on a 5-point Likert scale, with no statistically significant site differences but significant differences between low and high adherers high adherers indicating more willingness (r=.25, p=.05) (Supplementary Figure 3).

#### Adverse events

Additionally, adverse events were captured. 13 unrelated adverse events occurred. However, one participant reported hip pain during endurance exergames/walking on spot, which was categorized as a related adverse event. Additionally, 10 unrelated serious adverse events were recorded, with 5 being attributed to falls and 5 hospitalizations due to internal medical conditions.

### Usual care

Significant differences in physical and cognitive activities as well as therapies were observed between IG and CG for most items: participants in the CG spent significantly more time in sports, moderate physical activity, moderate cognitive activity, physical therapy, and cognitive therapy, whereas participants of the IG surpassed them only in the duration of intensive cognitive activity (Supplementary Table 2).

### Analysis of effectiveness

#### Training effects on physical, cognitive, and psychological outcomes

Table 4 shows differences and their effect sizes between pre- and post-assessments for each group separately and the interaction effect of group x time. A significant interaction effect was only found in the Go/No-Go test (Q(1, 53.17)=5.44, p=.02), and in Switzerland in the quality of life rating (Wald(1)= 5.86, p=.02).

**TABLE 4.** Pre-post comparisons and interaction effects in the secondary outcome measures.

| Variable | Site | IG (n=46) |  |  | CG(n=65) |  |  | Group x Time |
| --- | --- | --- | --- | --- | --- | --- | --- | --- |
|  |  | Pre (mean (SD)) | Post (mean (SD)) | Pre-Post | Pre (mean (SD)) | Post (mean (SD)) | Pre-Post |  |
| **RTT [RT in ms]** | All | 1129.48 (284.51) | 1108.44 (257.95) | $r=-.10, p=.32$ | 1174.88 (255.66) | 1252.37 (405.18) | $r=-.14, p=.11$ | $Q(1,53.37)=1.63, p=.21$ |
| | CH | 1008.58 (143.90) | 1060.81 (309.02) | $r=-.22, p=.38$ | 1098.97 (215.14) | 1040.38 (204.00) | $r=-.26, p=.20$ | $Q(1,9.61)=.40, p=.54$ |
| | IT | 1219.36 (329.06) | 1174.15 (263.01) | $r=-.17, p=.22$ | 1269.00 (164.77) | 1321.59 (278.67) | $r=-.14, p=.31$ | $Q(1,27.95)=.27, p=.61$ |
| | CY | 1031.02 (194.48) | 1011.38 (186.42) | $r=-.01, p=.94$ | 1123.37 (186.42) | 1281.42 (527.15) | $r=-.26, p=.05^*$ | $Q(1,15.24)=1.73, p=.21$ |
| **Go/No-Go [RT in ms]** | All | 977.63 (177.77) | 971.58 (167.31) | $r=.08, p=.42$ | 966.05 (162.82) | 1002.96 (188.23) | $r=.16, p=.07$ | **$Q(1,53.17)=5.44, p=.02^*$** |
| | CH | 964.57 (169.59) | 931.43 (171.62) | $r=.22, p=.38$ | 923.39 (148.70) | 927.68 (134.62) | $r=.05, p=.79$ | $Q(1,7.13)=2.21, p=.18$ |
| | IT | 1048.67 (156.46) | 1018.79 (169.65) | **$r=.30, p=.03^*$** | 1084.74 (112.57) | 1116.83 (136.72) | $r=.13, p=.35$ | $Q(1,27.96)=2.37, p=.14$ |
| | CY | 849.06 (155.69) | 905.50 (140.66) | $r=.28, p=.15$ | 878.37 (144.03) | 934.56 (175.88) | $r=.21, p=.12$ | $Q(1,14.46)=.09, p=.77$ |
| **Flexibility Test [RT in ms]** | All | 1663.89 (672.50) | 1721.74 (901.53) | $r=.01, p=.92$ | 1985.20 (1083.36) | 2032.48 (1533.85) | **$r=.17, p=.05^*$** | $Q(1,53.77)=.09, p=.76$ |
| | CH | 1348.58 (258.09) | 1186.95 (275.15) | **$r=-.67, p=.007^*$** | 1568.19 (488.77) | 1425.95 (397.51) | $r=-.29, p=.15$ | $Q(1,6.98)=.38, p=.56$ |
| | IT | 1888.07 (713.99) | 2033.94 (1091.30) | $r=-.02, p=.89$ | 2182.73 (949.97) | 2361.95 (1626.03) | $r=-.07, p=.60$ | $Q(1,24.57)=.19, p=.67$ |
| | CY | 1426.83 (636.94) | 1450.46 (385.13) | $r=-.04, p=.84$ | 1987.56 (1330.07) | 1998.24 (1702.07) | $r=-.19, p=.16$ | $Q(1,11.63)=.74, p=.41$ |
| **Sway path Length [mm]** | All | 458.89 (172.26) | 452.27 (182.49) | $r<.001, p=.099$ | 541.12 (528.11) | 552.63 (683.65) | $r=-.002, p=.97$ | $Q(1,53.33)<.001, p=.98$ |
| | CH | 440.04 (173.06) | 390.58 (199.05) | $r=-.23, p=.35$ | 456.33 (181.21) | 453.44 (132.85) | $r=-.05, p=.79$ | $Q(1,8.05)=.93, p=.36$ |
| | IT | 448.64 (139.67) | 489.74 (202.23) | $r=-.23, p=.10$ | 691.02 (813.63) | 743.48 (1071.50) | $r=-.11, p=.43$ | $Q(1,27.99)=.01, p=.92$ |
| | CY | 490.21 (230.79) | 418.18 (115.17) | $r=-.20, p=.31$ | 443.62 (159.45) | 424.73 (158.25) | $r=-.09, p=.48$ | $Q(1,15.95)=.02, p=.89$ |
| **Sway speed [mm/s]** | All | 15.29 (5.74) | 15.06 (6.08) | $r=-.004, p=.96$ | 18.05 (17.60) | 18.41 (22.79) | $r=-.009, p=.92$ | $Q(1,53.32)<.001, p=.99$ |
| | CH | 14.66 (5.78) | 13.00 (6.63) | $r=-.23, p=.35$ | 15.21 (6.05) | 15.12 (4.44) | $r=-.04, p=.84$ | $Q(1,8.02)=.95, p=.36$ |
| | IT | 14.95 (4.65) | 16.78 (6.73) | $r=-.22, p=.12$ | 23.07 (27.11) | 24.78 (35.71) | $r=-.10, p=.46$ | $Q(1,27.98)<.001, p=.99$ |
| | CY | 16.33 (7.70) | 13.92 (3.84) | $r=-.21$,<br>$p=.29$ | 14.78 (5.31) | 14.14 (5.28) | $r=-.09$,<br>$p=.48$ | Q(1,15.95)=.0<br>2, $p=.89$ |
| **CST -**<br>**path**<br>**devi-**<br>**ation**<br>**[mm]** | All | 2298.86<br>(2407.62) | 1860.13<br>(995.38) | $r=-.03$,<br>$p=.77$ | 1969.50<br>(1421.41) | 1925.22<br>(1209.81) | $r=-.04$,<br>$p=.66$ | Q(1,56.66)=.0<br>05, $p=.94$ |
| | CH | 2797.59<br>(1726.51) | 2007.33<br>(605.47) | $r=-.29$,<br>$p=.25$ | 2368.06<br>(1328.03) | 2140.80<br>(1409.43) | $r=-.10$,<br>$p=.62$ | Q(1,6.77)=.03,<br>$p=.87$ |
| | IT | 2044.18<br>(1265.22) | 2048.92<br>(1115.12) | $r=-.02$,<br>$p=.90$ | 2009.18<br>(1606.20) | 2242.70<br>(1260.30) | $r=-.10$,<br>$p=.46$ | Q(1,25.23)=1.<br>06, $p=.31$ |
| | CY | 2462.14<br>(4015.34) | 1421.00<br>(860.81) | $r=-.05$,<br>$p=.79$ | 1763.26<br>(1288.88) | 1549.40<br>(993.84) | $r=-.13$,<br>$p=.33$ | Q(1,10.26)=.0<br>2, $p=.90$ |
| **CST –**<br>**com-**<br>**plete-**<br>**ness**<br>**[%]** | All | 42.29<br>(23.39) | 38.76<br>(24.22) | $r=-.14$,<br>$p=.18$ | 38.29<br>(24.30) | 37.36<br>(22.05) | $r=-.01$,<br>$p=.89$ | Q(1,54.00)=.0<br>4, $p=.85$ |
| | CH | 41.64<br>(19.36) | 42.71<br>(18.69) | $r=-.15$,<br>$p=.55$ | 41.61<br>(19.71) | 37.03<br>(22.71) | $r=-.10$,<br>$p=.52$ | Q(1,9.99)=.29,<br>$p=.60$ |
| | IT | 39.39<br>(25.86) | 32.83<br>(26.55) | $r=-.26$,<br>$p=.07$ | 26.48<br>(20.34) | 25.10<br>(15.35) | $r=-.02$,<br>$p=.88$ | Q(1,28.00)=.8<br>3, $p=.37$ |
| | CY | 48.28<br>(20.94) | 47.73<br>(20.379) | $r<.001$,<br>$p=.99$ | 47.41<br>(25.55) | 48.46<br>(21.59) | $r=-.03$,<br>$p=.82$ | Q(1,15.94)=.0<br>1, $p=.92$ |
| **STS**<br>**[n full**<br>**stand**<br>**ups]** | All | 12.16 (4.96) | 12.56 (5.14) | $r=-.06$,<br>$p=.55$ | 13.34 (4.19) | 13.23 (4.33) | $r=-.04$,<br>$p=.69$ | Q(1,63.92)=.0<br>07, $p=.93$ |
| | CH | 12.00 (5.45) | 14.13 (4.61) | $r=-.48$,<br>$p=.06$ | 12.83 (3.90) | 13.50 (3.73) | $r=-.23$,<br>$p=.25$ | Q(1,8.47)=1.4<br>2, $p=.27$ |
| | IT | 10.71 (5.38) | 11.58 (6.31) | $r=-.13$,<br>$p=.37$ | 12.36 (4.52) | 12.04 (4.81) | $r=-.09$,<br>$p=.51$ | Q(1,27.85)=.0<br>4, $p=.84$ |
| | CY | 14.92 (2.25) | 13.38 (1.94) | $r=.33$,<br>$p=.10$ | 14.48 (3.87) | 14.22 (3.97) | $r=-.09$,<br>$p=.51$ | Q(1,14.34)=4.<br>33, $p=.06$ |
| **TUG**<br>**[s]** | All | 9.38 (4.98) | 9.50 (4.48) | $r=-.04$,<br>$p=.68$ | 7.93 (3.49) | 8.56 (3.59) | $r=-.14$,<br>$p=.12$ | Q(1,40.35)=.3<br>1, $p=.58$ |
| | CH | 10.11 (8.38) | 8.57 (6.51) | $r=-.52$,<br>$p=.04^*$ | 7.82 (3.40) | 7.30 (2.30) | $r=-.10$,<br>$p=.61$ | Q(1,9.65)=.96,<br>$p=.35$ |
| | IT | 10.84 (3.67) | 10.47 (4.67) | $r=-.22$,<br>$p=.12$ | 9.93 (3.98) | 10.42 (4.69) | $r=-.05$,<br>$p=.72$ | Q(1,24.14)=.8<br>0, $p=.38$ |
| | CY | 6.11 (2.93) | 8.22 (1.59) | $r=.46$,<br>$p=.02^*$ | 6.20 (1.81) | 7.43 (1.88) | $r=-.28$,<br>$p=.04^*$ | Q(1,12.99)=2.<br>85, $p=.12$ |
| **TUG-DT**<br>**[s]** | All | 12.70 (7.27) | 12.65 (7.17) | $r=-.06$,<br>$p=.59$ | 10.44 (4.86) | 11.23 (5.10) | $r=-.07$,<br>$p=.40$ | Q(1,36.28)<.0<br>01, $p=.98$ |
| | CH | 11.08 (9.20) | 10.26 (7.56) | $r=-.18$,<br>$p=.46$ | 10.48 (5.93) | 9.03 (3.17) | $r=-.26$,<br>$p=.20$ | Q(1,6.68)=.06,<br>$p=.81$ |
| | IT | 15.43 (6.65) | 14.26 (8.21) | $r=-.26$,<br>$p=.07$ | 13.00 (5.01) | 13.07<br>(6.07) | $r=-.20$,<br>$p=.16$ | Q(1,27.55)=1.<br>14, $p=.30$ |
| | CY | 8.43<br>(4.80) | 11.04 (3.53) | $r=-.26$,<br>$p=.07$ | 8.13 (2.81) | 10.54 (4.37) | $r=-.39$,<br>$p=.003^*$ | Q(1,10.21)=1.<br>41, $p=.26$ |
| **TUG DT**<br>**cost**<br>**[%]** | All | -33.62<br>(26.37) | -31.34<br>(34.06) | $r=-.10$,<br>$p=.33$ | -32.43<br>(27.58) | 31.34<br>(27.24) | $r=-.07$,<br>$p=.42$ | Q(1,48.58)=.5<br>6, $p=.46$ |
| | CH | -10.10<br>(12.27) | -23.98<br>(41.93) | $r=-.25$,<br>$p=.31$ | -31.99<br>(34.32) | -23.70<br>(22.67) | $r=-.23$,<br>$p=.27$ | Q(1,8.95)=.84,<br>$p=.38$ |
| | IT | -40.39<br>(29.64) | -33.95<br>(36.12) | $r=-.22$,<br>$p=.11$ | -33.07<br>(20.94) | -25.37<br>(18.55) | $r=-.24$,<br>$p=.09$ | Q(1,23.81)=.0<br>4, $p=.85$ |
| | CY | -35.09<br>(17.17) | -32.38<br>(25.80) | $r=-.05$,<br>$p=.79$ | -32.04<br>(30.53) | -39.94<br>(33.29) | $r=-.14$,<br>$p=.29$ | Q(1,13.81)=.2<br>4, $p=.15$ |
| **ABC**<br>**[score]** | All | 80.13<br>(18.42) | 77.70<br>(19.23) | $r=-.15$,<br>$p=.14$ | 85.43<br>(15.89) | 83.24<br>(15.75) | $r=.21$,<br>$p=.02^*$ | Q(1,57.57)=.0<br>03, $p=.96$ |
| | CH | 85.96<br>(16.06) | 89.41<br>(14.89) | $r=-.44$,<br>$p=.08$ | 87.09<br>(9.04) | 86.31<br>(12.47) | $r=-.16$,<br>$p=.41$ | Q(1,9.53)=.79,<br>$p=.39$ |
| | IT | 71.40<br>(19.02) | 70.31<br>(20.20) | $r=-.17$,<br>$p=.24$ | 78.76<br>(20.26) | 76.35<br>(20.26) | $r=-.21$,<br>$p=.14$ | Q(1,26.58)=.0<br>8, $p=.77$ |
| | CY | 93.34<br>(5.25) | 84.70<br>(14.51) | $r=-.38$,<br>$p=.05^*$ | 90.42<br>(11.36) | 87.83<br>(9.92) | $r=-.24$,<br>$p=.08$ | Q(1,11.09)=.1<br>8, $p=.68$ |
| **Quality**<br>**of life** | All | 3.78<br>(.89) | 3.81<br>(.97) | $r<.001$,<br>$p=.99$ | 4.05<br>(.89) | 3.97<br>(.82) | $r=-.10$,<br>$p=.30$ | Wald(1)=1.57,<br>$p=.21$ |
| | CH | 4.38<br>(.52) | 4.5<br>(.53) | $r<.001$,<br>$p=.99$ | 4.5<br>(.52) | 4.17<br>(.39) | $r=-.37$,<br>$p=.07$ | **Wald(1)=5.86,**<br>**$p=.02^*$** |
| | IT | 3.40<br>(.96) | 3.39<br>(1.12) | $r=-.02$,<br>$p=.90$ | 3.46<br>(1.02) | 3.46<br>(0.88) | $r=-.03$<br>$p=.82$ | Wald(1)=.05,<br>$p=.82$ |
| | CY | 4.15<br>(.55) | 4.14<br>(.38) | $r<.001$,<br>$p=.99$ | 4.39<br>(.57) | 4.36<br>(.68) | $r=-.04$,<br>$p=.79$ | Wald(1)=.01,<br>$p=.94$ |
| **Satis-**<br>**faction**<br>**with**<br>**health** | All | 3.57<br>(.98) | 3.48<br>(1.00) | $r=-.09$,<br>$p=.36$ | 3.81<br>(.82) | 3.81<br>(.80) | $r=-.02$,<br>$p=.79$ | Wald(1)=.46,<br>$p=.50$ |
| | CH | 3.75<br>(1.04) | 4.00<br>(.53) | $r=-.14$,<br>$p=.59$ | 4.00<br>(.85) | 3.67<br>(.65) | $r=-.04$,<br>$p=.07$ | Wald(1)=2.99,<br>$P=.08$ |
| | IT | 3.20<br>(1.04) | 2.96<br>(1.07) | $r=-.19$,<br>$p=.19$ | 3.46<br>(0.93) | 3.54<br>(.98) | $r=-.01$,<br>$p=.97$ | Wald(1)=1.21,<br>$p=.27$ |
| | CY | 4.15<br>(.38) | 4.08<br>(.49) | $r=-.06$,<br>$p=.77$ | 4.04<br>(.58) | 4.11<br>(.57) | $r=-.06$,<br>$p=.62$ | Wald(1)=.55,<br>$p=.46$ |

Note. ABC: Activities-specific Balance Confidence; CH: Switzerland; CST: Coordinated Stability Test; CY: Cyprus; DT: dual-task; IT: Italy; Q: Q-statistic (degrees of freedom with-groups, degrees of freedom between group); $r$ =Pearson correlation coefficient $r$ ; RT: Reaction Time; RTT: Reaction Time Test; SD: Standard deviation; STS: Sit-to-Stand Test; TUG: Timed up-and-go; Wald(x): Wald statistic (degrees of freedom)
\* $p < 0.05$ , bold values indicate significance.

As additional explanatory analyses, which were not planned in the protocol, within-group differences were investigated. In the IG, significant differences could only be detected when analyzing the three study sites separately. Thus, IG participants from Switzerland (CH) improved significantly in the Flexibility test (r=-.67, p=.007), and in the TUG Test (r=-.52, p=.04), and participants from Italy in the Go/No-Go Test (r=.30, p=.03). However, in participants from Cyprus, the TUG performance as well as the subjective balance confidence declined significantly (r=.46, p=.04; r=-.38, p=.05). In CG participants, the subjective balance confidence of all participants from all countries declined (r=.21, p=.02) as well as the reaction time in the Flexibility test (r=.17, p=.05). CG participants from Cyprus furthermore worsened in the RTT (r=.26, p=.05), the TUG (r=.28, p=.04), and the TUG DT (r=-.39, p=.003).

#### Intention-to-treat (ITT) analysis

The results of the per protocol analysis and the intention-to-treat analysis did not show any differences. Thus, in the whole study population, the only interaction effect was found in the Go/No-Go test (Q(1,70.00)=5.86, p=0.02).

## Discussion

The primary aim of this pragmatic RCT was to evaluate the feasibility of a home-based exergame training in older adults. Additionally, we investigated potential training-related changes in physical, cognitive, and psychological functions. In brief, the training was feasible and safe as indicated by high levels of adherence and enjoyment, alongside minimal attrition rates and perceived exertion/workload. However, significant interaction effects were observed solely for inhibition, and significant within-group differences were only detected in certain functions at certain study sites.

### Feasibility of the training program

#### Attrition, adherence, and enjoyment

The attrition rate of 11.8% was close to the threshold of 10% which was defined as a threshold for an acceptable feasibility by other studies [35], and well below the limit of 20% we set prior to the study start [21]. Similarly, the mean adherence rate exceeded the threshold of 70% which has been defined as “being adherent to the training. Based on these findings the home-based exergame training is deemed feasible.

Looking at previous studies, the attrition rate of this study was similar to [36]–[38] or even lower than [9], [39]–[41] rates reported in other comparable unsupervised home-based exergame interventions. Likewise, the high adherence rates are also in line with previous comparable inpatient [6], [42] as well as home-based exergame intervention studies [8], [43]–[46] and are further confirmed by a systematic review indicating higher adherence to technology-based interventions compared to traditional interventions regardless of whether they are supervised or unsupervised [16]. One explanation might be that in this study as well as in previous technology-based intervention studies, adherence was automatically recorded which might have resulted in a higher motivation or discipline, respectively, to adhere to the training program. Another explanation is provided by the authors of the aforementioned review who concluded that the high adherence rates can largely be explained by high enjoyment associated with technology-based interventions [16]. The latter, however, is not completely supported by the findings of the current study. Despite the overall high enjoyment levels, as anticipated considering the “gamification” aspect [14] and consistent with previous exergame studies [6], [20], [42], [47], [48], surprisingly, low adherers reported higher enjoyment compared to high adherers. Moreover, explanatory analyses showed no correlations between adherence and enjoyment at any study site.

Upon closer examination of the characteristics of participants from Cyprus and Italy, where the disparity between enjoyment and adherence was most pronounced, a possible explanation emerged. Participants from Cyprus had the lowest adherence rates despite having less comorbidities, fewer falls, the highest enjoyment, and the lowest frustration score (NASA-TLX) compared to the other study sites. Conversely, in Italy, participants, despite being overall more frail (as indicated by their demographics and baseline measures (Supplementary table 3), reporting the highest training load, lowest balance confidence, and least enjoyment, still had high adherence rates. This finding might be explained by the Health Belief Model stating that health behavior is dependent on two factors: (1) threat perception (including perceived susceptibility to health problems and perceived severity), and (2) behavioral evaluation (including perceived benefits and barriers) [49]. This aligns also with a prior focus group study investigating older adultś views on the exergame device used in the present study which elucidated that personal benefits and meaningful purposes play pivotal roles in technology adoption. [19]. To sum up, the good health conditions in Cyprus may have reduced the perceived need for training despite high enjoyment, suggesting that adherence is influenced by health status rather than enjoyment alone. This, in turn, would indicate that a poor health might increase adherence, but at the same time possibly limits enjoyment due to for instance higher perceived demands, and a low balance confidence as apparent in Italy. Hence, in the current study population, enjoyment may not be directly linked to adherence but rather indirectly through the overall health status.

However, it should also be taken into consideration that older adults from Cyprus were less educated compared to participants from the other study sites (Supplementary Table 1) and have demonstrated a lower ICT competency likely attributable to limited financial resources as well as a lower acceptance of technology [20]. This, in turn, may also have contributed to the lower adherence rates. Conversely, the novelty value of the COCARE system may have heightened the enjoyment levels among participants from Cyprus.

This might also elucidate the high levels of both adherence and enjoyment in Switzerland. A previous usability study revealed that participants from Switzerland had higher acceptance ratings of the COCARE system compared to the other study sites [20], which likely contributed to a higher adherence and higher enjoyment. Moreover, it is noteworthy, that more than half of the participants from Switzerland lived alone, potentially having more time available to engage with the training program.

Nevertheless, it must also be considered that the intervention period was relatively short which is why the interplay between enjoyment and adherence must further be explored in future longitudinal studies. Still, enjoyment remains a crucial factor for feasibility, since it has shown to be closely linked to intrinsic motivation [50] and likely contributes more to long-term adherence for which it appears to be especially important at the beginning of an intervention [51]. Therefore, the overall high enjoyment scores are an important finding supporting the feasibility of the investigated exergame training.

#### Perceived workload

The mean NASA-TLX raw score of 40.1 points observed in our study falls significantly below mean values reported in a review by Grier et al. (2015) who found that in previous studies using the NASA-TLX, the average load of physical activities was 62.0, of cognitive tasks 46.0, and of video games 56.5 [52]. This suggests that especially the physical component of the step-based exergame training was notably less demanding compared to other physical activities and that no overexertion occurred.

Conversely, regarding the cognitive component, the total raw score approached those of other cognitive activities and considering the higher scores on the item “mental load” it appears that the exergames imposed greater cognitive demands than physical ones. This aligns with earlier investigations using the Dividat Senso [6], [42] and is not surprising considering that most games target various high-order cognitive functions. Furthermore, physically rather healthy older adults had mixed evaluations concerning the physical demands [19], [20] supporting the findings of this study.

Overall, the low training load is most likely accountable for participants expressing a wish for additional and more entertaining games and for some participants dropping out due to lack of time and interest. Consequently, for cognitively and physically high functioning older adults, adjustments to the training plan seem necessary, such as incorporating more games per level and/or modifying the progression rules to allow for faster progression even without the occurrence of a performance plateau or using additional equipment in order to make the motor task more challenging [3]. Generally, adding more games would allow even more personalization and thereby possibly more motivation and no decline in adherence rates over time. However, the responses may differ among low-functioning older adults, and it is noteworthy that the combination of a low frustration score and high enjoyment scores suggests that, for some participants, the training intensity remained satisfactory.

#### Help requests

Another important measure of feasibility was the help requests posed by participants during the unsupervised training period. Despite the relatively large sample size, help requests were infrequent (15 help requests within 12 months data collection period), which is possibly also reflected in the low frustration score of the NASA-TLX questionnaire. Nevertheless, these requests must be considered for further system development, particularly the criticisms of the mat’s sensitivity, which has been a recurring issue across different iterations of the system [20], [46]. Surprisingly, contrary to expectations based on the previous studies [20], [46], only a single participant required assistance in understanding the instructions despite the regular introduction of new games. This supports the usefulness of the familiarization phase prior to the independent training at home. Overall, these findings are another indicator of a good feasibility of the home-based exergame training program - given a frequent availability of human support.

#### Willingness to continue

The positive outcomes regarding the feasibility of the training are mirrored in the high agreement ratings concerning the willingness to continue a similar home-based exergame training. Furthermore, the absence of site differences suggests that this is applicable for a broad spectrum of older adults with diverse characteristics. The significant difference between low and high adherers, with low adherers expressing less willingness to continue the training program despite higher enjoyment, supports the notion that enjoyment is not the (only) determining factor when it comes to the acceptance of the training program and system.

### Effectiveness of the training program

#### Training effects on cognitive functions

The results showed significant time x group interaction effects in favor of the intervention group only for response inhibition as measured by the Go/No-Go test. Furthermore, in Italy, the intervention group improved significantly in this test with a medium effect size. However, pre-post comparisons of the whole sample show only small within-group effect sizes in both IG and CG. These outcomes are overall consistent with previous literature [9], [39], [44] which reported significant effects of various in-home exergame training on inhibition with effect sizes ranging from small to medium. Conversely, other similar studies found no significant effects on inhibition [37], [43].

No significant interaction effects were detected for the other cognitive functions (mental flexibility and processing speed/reaction time). This is in line with previous comparable studies which did not find any effects on similar cognitive functions [39], [43], [53], but contrasts with other earlier studies demonstrating significant effects of home-based exergames on choice stepping reaction time [36], [37], processing speed [44], and attention [37].

Comparing our study with those demonstrating positive effects on cognitive functions, two major differences and therefore potential explanations for the limited effects emerge: (1) Previous studies had longer training duration and/or higher training volumes. This is supported by reviews recommending exercise sessions lasting at least 30 minutes, 1-3 times per week for a minimum of 12 weeks [4] or even up to 6 months [54] to enhance cognitive functions. (2) Other exergame training programs incorporated more exhaustive physical training components which have shown to have transfer effects on cognition. Thus, it is likely that in our study, the training dose/intensity as well as the physical component were not sufficiently high, confirming the perceptions of the participants described above. On the other hand, Sturnieks et al. (2024) [43] conducted a home-based stepping exergame intervention over 6 months with a training volume of 120 min per week, including a huge number of participants, and yet did not observe any significant effects on cognitive functions either. This raises an additional explanation: in both their study and ours, participants were physically and cognitively rather high functioning and, as Sturnieks et al. (2004) [43] pointed out, the “*divergence of results may also reflect the current study being undertaken in a more able and healthy group, in which changes in function are harder to detect”.* This assumption is supported when examining the different trial sites separately. Notably, participants from Italy improved significantly in the Go/No-go test with a medium effect size while having the lowest baseline-performance in all cognitive assessments.

Finally, it must be acknowledged that the control group underwent significantly longer cognitive therapy sessions as part of their usual care programs and still experienced significant declines in the Go/No-go test, the Flexibility test and partly in the RTT (in Cyprus). In contrast, participants from the IG were able to maintain their level of functioning, which, although not the aim of the intervention, could still be considered a small achievement, particularly in older adults.

#### Training effects on physical functions

No significant interaction effect on any physical function was found for the whole study population. Only in the TUG test, participants from Switzerland improved significantly, whereas the performance of those from Cyprus decreased significantly.

Overall, these findings again align with the aforementioned study of Sturnieks et al. [43] who did not find any physical effects of their step-based exergame training. However, most similar home-based exergame studies could find significant improvements in the intervention groups in the TUG [38], [40], [44], [55] and significant interaction effects in the TUG-DT [36], [37], 30s STS [9], and sway path length [36]. Coordinated Stability was only examined in two similar studies which did not observe significant effects [43], [53].

Two major differences between those studies demonstrating effects compared to our study are minutes [55] and most also had longer intervention periods between 12 [38], [40] and 16 weeks [9]. Furthermore, different exergame devices were utilized in these studies, such as the Microsoft Kinect motion capture system [38], [40], Xbox Kinect and Wii Fit Plus [44], Nintendo game systems [55], and IMU-based motion detection systems. These devices allow for both upper and lower limb movements, enabling exercises based on more complex movements with more degrees of freedom and higher intensities. Such exercises included aerobic exercises and dance movements [9], [38], [40], yoga [55], and strength exercises [38], [40], [55].

Thus, as already indicated by the subjective workload ratings, a potential explanation for missing physical effects in our study is that the step-based training program may not be sufficiently physically challenging for high functioning older adults. This could be attributed to three factors. First, compared to previous studies, the training volume was lower. Second, games with clear focus on physical functions were underrepresented compared to cognitive games, and third, stepping movements might not be demanding enough in this population. Moreover, the two games designed to target endurance, “Rocket” and “Flaneur”, are based on walking on spot, which might not have direct transfer effects on forward walking as required in the TUG. Additionally, these two games were less favored by participants who preferred games with a clear aim.

Regarding the TUG, additional analyses revealed that participants whose TUG performance even decreased, had significantly better baseline performances (p<.001) and less comorbidities (p=.002) (and lower MMSE, p=.048) compared to the other participants. Besides, at T3, their TUG completion times were still lower compared to those who improved – though not statistically significantly - and also below reference values [56]. Another point to consider is that the whole study population had average baseline TUG times well below those indicative of impaired physical functioning [57]. Taken together, these observations indicate that a ceiling effect might have occurred and/or participants were less motivated to train physical functions due to their already good physical functioning – again as suggested by the aforementioned Health Belief Model [49].

Concerning the training effects on balance, the findings were unexpected since some games directly targeted balance. However, according to previous research, balance is not a general category or ability, respectively. Rather, 6 distinct components of balance have been identified [58] and evidence suggests a high degree of specificity of balance training. Thus, balance training enhances the performance solely in the targeted component, without eliciting transfer effects onto another balance components [59]. In this study, most balance exergames were based on weight shifting and, thereby, targeted only static balance, while the stepping exergames trained dynamic stability, requiring a change in the base of support, and anticipatory postural control [58]. Consequently, no effects on functional stability and functional balance, as assessed through the Coordinated Stability Test and the TUG test could have been expected.

Furthermore, two more issues warrant consideration: Problems with mat sensitivity occurred primarily during balance games which might have led to frustrations and reduced engagement and future studies should consider alternative Coordinated Stability parameters, such as an error value which was for instance used in a study by Gschwind et al. (2015) [53].

#### Training effects on balance confidence

No significant intervention effect on balance confidence was observed in our study, while previous studies investigating stepping exergames or balance board training present inconsistent results [60]. In our study, the lack of effect may again be attributed to a ceiling effect since the overall pre-intervention ABC score was 80.13 with participants in Cyprus even reaching 93.34 points and according to Myers et al. [61], scores above 80 on the ABC scale are indicative of highly functioning and physically active older adults, suggesting limited room for further enhancement of balance confidence. Correspondingly, according to Büla et al. (2011) [62] individuals with a low balance confidence or those at risk of falls are more likely to respond to interventions. Additionally, they found that most effective interventions for improving balance confidence in community-dwelling older adults are multifactorial, incorporating physical exercise components, and that improvements are mediated by improvements in gait and balance. The latter suggests that ABC scores declined alongside the decreased TUG performance, and it appears that the interventions’ effectiveness might have been compromised again by its low physical intensity, particularly in high functioning older adults.

#### Training effects on quality of life

No significant intervention-related effects on quality of life and satisfaction with health were evident in the whole sample. Only in the Swiss subgroup, an interaction effect was found. This is consistent with previous literature showing mixed results [11]. Cacciata et al.(2019) [11] emphasized that physical exercise related changes in quality of life require high intensities, yet the determination of appropriate exergame intensities requires further investigation. Finally, baseline values in both groups were notably high in our study, potentially leading to a ceiling effect which may have occurred alongside a possible ceiling effects in balance confidence which has been suggested to impact quality of life [63].

## Limitations

A major limitation is that in Switzerland, the targeted sample size could not be reached. Numerous interested participants did not meet the inclusion criterion of a prescription for rehabilitation. This might be due to the fact that in Switzerland, exercise therapies are primarily offered as part of rehabilitation programs and are still rarely used as a way of preventing physical function decline or as part of long-term follow-up care [64]. The different sample sizes might then have contributed to the various significant differences between trial sites.

Furthermore, despite including only individuals with a prescription for rehabilitation, most included participants still turned out to be high functioning which is the major reason for which the proposed training plan did not sufficiently match the functional abilities of participants. On the other hand, since this is a feasibility study, safety concerns were in the focus and the fact that no adverse events occurred provides a good safety basis for increasing the intensity or conduct the same training program with frailer participants in future RCTs.

Moreover, neither the participants nor the assessors were blinded for reasons elaborated above. However, this is not required in pragmatic RCTs and allowed to better concentrate on feasibility measures. Generally, it must be considered that the study was designed as a pragmatic RCT which is recommended for the evaluation of complex intervention. This leads to a limited internal validity but at the same time to a high external validity since real-world evidence is provided allowing generalizability to many real-world settings.

Furthermore, in future studies, standardized questionnaires for the assessment of quality of life and satisfaction with health should be used which are more comprehensive and sensitive to change.

Finally, IG and CG participants differed in several usual care variables which were not included in the robust mixed ANOVAs since an excessive number of covariates increases the model complexity and can make the interpretation of the results more difficult. In addition, there is a risk of multicollinearity. This might partly explain missing interaction effects but does not explain missing within-group effects.

## Conclusion

The step-based exergame intervention using the Dividat Senso Flex in home-setting appeared to be feasible and safe which was shown through low attrition rates, high adherence rates, high enjoyment, a high willingness to continue the training program, and a low number of help requests. This supports its application in out-patient rehabilitation settings. Additionally, it was effective in improving inhibition. However, to improve further cognitive, physical, and psychological functions as well as quality of life in non-acute, rather high functioning older adults, it appears necessary to increase the training load by adding more games and by adapting the progression speed.

## Data Availability

All data is publicly available on Zenodo (https://doi.org/10.5281/zenodo.11105179).

## Acknowledgments

The authors would like to thank VAMED Rehabilitation Centre and Atelier 1 for their support in recruitment and for providing rooms and equipment for the supervised training in Switzerland. Also, we would like to thank all study participants. Finally, we would like to thank Sarina Ifanger, Amilen Souto-Cortes, Lisa Lutz, Sandro Roth, and Jessica Staubli for their support in data collection and training supervision.

This work was funded by the European Union and the involved national funding authorities (Innosuisse, the Swiss Innovation Agency; the Italian ministry of health; and the Cyprus Research and Innovation Foundation) as part of the AAL Association joint program (aal-2020-7-145-CP). In Italy, the study was also supported by Ricerca Corrente 2023-2024 (Italian Ministry of Health).

## Author contributions

JS and EG conceived and designed the study and JS, EK, ES, DK, DDB, FR, IC, JR, LL, MF, RV, and SM performed data collection. JS undertook all statistical analysis and wrote the original draft.EG and EDB acted as supervisors. All authors critically revised the manuscript for content and approved the version submitted for publication, and they agree to be accountable for all aspects of the work.

